# SARS-CoV-2 infection among patients with multiple sclerosis; A cross-sectional study

**DOI:** 10.1101/2020.10.17.20214429

**Authors:** Mahnaz Bayat, Alireza Fayyazpoor, Afshin Borhani Haghighi, Daniyal Salehi, Hossein Molavi Vardanjan, Maryam Poursadeghfard

**Author notes:** **Corresponding author:** Maryam Poursadeghfard, Assistant Professor in neurology, Clinical neurology research center, Shiraz University of Medical Sciences, Shiraz, Iran, **E-mail:**, **Postal Code :** 71365-1689.

## Abstract

**Background:** Neurological disability associated with multiple sclerosis and immunosuppressive or immunomodulatory therapy which is administered for it may increases the risk of SARS-CoV-2 infection and its morbidity/mortality.

**Objective:** In this study, we evaluated the infection rate and the severity of SARS-CoV-2 infection in patients with multiple sclerosis (MS)

**Methods:** One thousand and three hundred and sixty one MS patients from Fars province, south of Iran, were interviewed by phone from April 3 to June 20, 2020. Basic demographic data, information about MS disease and any symptoms or laboratory results relevant to COVID-19 were gathered and reviewed by treating neurologist and MS nurses. SPSS version 22 was used for data analysis.

**Results:** 68 (5%) of MS patients were suspected cases and 8 (0.58%) of all patients with positive real-time reverse transcription polymerase chain reaction (RT-PCR) or chest CT were in the confirmed group. 5 cases of the confirmed group needed hospitalization. Two patients died while both of them had PPMS and were taking rituximab. The frequency rate of suspected cases with RRMS was 57 (87.7%), followed by PPMS 5 (7.7%) and CIS 2(3.1%). In the confirmed group 37.5% had RRMS, 50% had PPMS, 25% use corticosteroid drug, and 50% were on rituximab. 62.5% of confirmed cases had high disability level and need assistance to walk. 36.8% of suspected and 25% of the confirmed cases were on IFN-β1; eventually all of them recovered well from COVID-19 infection.

**Conclusion:** The present study showed that rate of developing COVID-19 in MS patients are similar to the general population and the frequency of PPMS phenotype, rituximab therapy and corticosteroid therapy were higher in the confirmed group.

## Introduction

The managing of patients who need to receive immunosuppressive or immunomodulatory drugs rapidly has to change during the current coronavirus disease 2019 (COVID-19) pandemic. Multiple sclerosis (MS) is a chronic autoimmune inflammatory disorder in which the immune system attacks the myelin and by disruption of neuronal signal propagation, causes progressive disabilities (1). Approximately up to 70% of people with MS receive disease-modifying therapies (DMTs) that influence the immune response. There are also evidences that treatment with second lines of DMTs significantly increase the incidence rate of infection in MS patients compare to general population (2-4). It seems immunosuppression therapy combined with neurological disability in MS increases the risk of severe COVID-19 disease and associated death (5). Treatment with type 1 interferons in MS patients has showed a lower risk of pneumonia (3) and a few published studies have supported the role of type 1 interferons in the potential therapy of COVID-19 infection (6-9). Hence, evaluating not only the risk of Covid-19 but also severity of the infection in MS patients is an important issue in global healthcare system. Therefore, the demand for data to describe the frequency, morbidity, mortality and in a word, the evolution of COVID-19 infection in MS patients grows rapidly around the world (10). Some studies have been reported the severity and outcomes of COVID-19 infection in MS patients (11-14). However, few epidemiological studies with high number of population are now available; accordingly, we think larger and better-characterized data in population-based studies with sufficient sample size are needed for better interpretation of the results. Here, we designed our cross-sectional study on a large group of PwMS to pick up cases with confirmed or suspected COVID-19 infection and evaluate the effects of that on this particular population compared with normal people. The aim of this cross-sectional study was to determine the frequency rate and severity of disease in MS patients of southern Iran. Due to the fact that, Iran is a country with high prevalence of MS which is rising over time (15) and is high in the number of infection rate or deaths due to COVID-19 (16, 17) it is hoped that the obtained results from this population-based study will contribute to a deeper understanding of frequency and severity of COVID-19 in MS patients with DMT.

## Methods

This Cross-Sectional Study was approved by Iran National Committee for Ethics in Biomedical Research with the registration number IR.SUMS.REC.1399.26. 1700 patients were contacted by phone from April 3 to June 20, 2020 and 1361 patients agreed to participate in our study. All patients were from Fars province, one of the most affected areas of Iran by covid-19 pandemic. Patients or caregivers answered our checklist used in this study.

Data elements included in age, sex, city, job, types of MS, types of current DMT (or no DMT), duration of current DMT, information about comorbidities, history of medications other than DMT, disability level by a self-defined scoring scale (score =1-6), a history of close contact with a confirmed COVID-19 case or with someone who has respiratory symptoms during the past 14 days. Additional information was also taken into account including (a) Confirmed infection by positive real-time reverse transcription polymerase chain reaction (rRT-PCR) checked by oropharyngeal swab or a positive chest computed tomographic (CT) scan, (b) Suspected infection by showing one of the clinical signs of covid-19 infection after February 20, 2020: such as cough, fever, shortness of breath, loss of taste or smell or one of these signs plus another symptom such as sore throat, dyspnea, body pain, headache, muscle pain, or diarrhea, (c) Severity of disease by duration of symptoms, referral or non-referral to a medical specialist, hospitalization (ward or intensive care unit [ICU]), and patient death.

We considered MS patients in three case groups: 1-Suspected COVID-19 case (any patients meeting the clinical symptoms without diagnostic imaging or laboratory criteria (rRT-PCR)) 2-Confirmed COVID-19 case (any patients meeting the clinical symptoms plus confirmed rRT-PCR- or a positive (CT) scan). 3-Healthy case (any patients without any clinical symptoms of COVID-19 infection). We estimated the occupational risk factors of MS patients for covid-19 infection based on the front line was published on April 15, 2020 by Marcus Lu (18). The patient’s job categorized into three groups: low, intermediate, and high risk.

Because we had to determine the degree of disability of patients by phone, it was not possible to use the EDSS scoring. We categorized the MS patients according to self-defined scoring to following groups: 1 = Able to walk more than 500 meters without aid, 2= Able to walk less than 500 meters, 3= Restricted to stick, 4= Restricted to walker, 5= Restricted to wheelchair, 6= Restricted to bed. MS patients with a score three or higher, which require assistance to walk, were considered in another group.

Statistical package for social sciences (SPSS Inc., Chicago, IL, USA) version 22 was used for data analysis. P < 0.05 was considered statistically significant. One-way ANOVA and Chi-square analysis was performed for detection of significant differences in the clinical characteristics, demographic, and the frequency of COVID-19 symptoms between healthy, suspected and confirmed cases.

## Results

1361 MS patients participated in this research (Table 1). 77% of all patients were female with mean age (38.48 ± 9.40). 1171 (86.6%) of patients were from Shiraz and 190(14 %) from other provincial cities. Most patients (95.1%) had job with low risk of COVID-19 and 1.6% had high-risk job based on described in the method. 85.5% of MS patients had RRMS and 87.6 % of the all respondents received one of the DMTs. The frequency of prescribed DMTs were as follow: rituximab 27.7%, IFN-β 26.9%, fingolimod 12.3%, dimethyl fumarate (DMF) 9.0%, GA 7.2%, teriflunomide 1.8%, natalizumab 1.2% and 74.5% of patients were treated for more than three months. The most frequency of comorbidity in MS patients was hypothyroid disease, and then second and third ranks belong to hypertension and cardio vascular diseases (CVD), which were presented in 6.31%, 4%, and 3% of patients respectively. The frequency of prescribed drugs other than DMT as followed: levothyroxine 4.84%, corticosteroid 3.7%, antihypertensive 3.8% .89.8% of all respondents had disability scale less than 3. These patients do not require assistance to walk.

The following flowchart shows the different groups of MS patients with rate of mild symptoms and hospitalization after COVID-19 infection.

Out of 1361 patients (Fig. 1), 68 (5%) were suspected cases, these patients were qualified by their reported at least two defining symptoms of COVID-19 infection and 8 (0.58%) of all MS patients with positive RT-PCR or chest CT were in confirmed group. 40 out of 68 patients improved by staying home without medical attention. 28 patients offered to the physician due to their pulmonary symptoms and 21 were recommended to stay home and monitor the severity of symptoms and 7 patients showed severe clinical symptoms that required hospital admission with negative test and were discharged after 3 days of hospitalization. 3 cases had positive PCR test without hospitalization however, 5 cases had positive RT-PCR or CT results needed hospitalization due to the severity of pulmonary symptoms and shortness of breath, 3 of them did not require ICU care or intubation and were eventually discharged from the hospital. In addition, 2 of the confirmed cases died while both of them were female, had PPMS with relatively high disability score; one was bed-restricted and the other able to walk without assistance less than 500 meters. Their DMTs were rituximab and (comorbidity)

**Figure 1:**
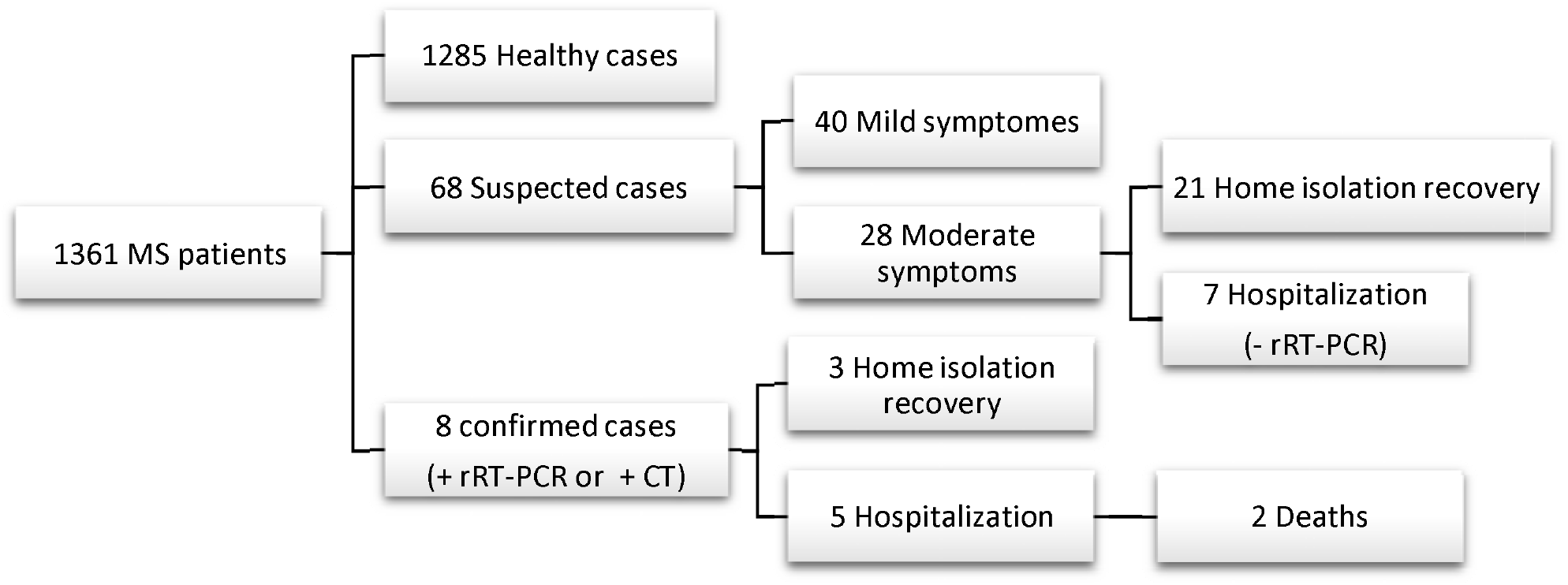
Flowchart of the study population

The demographic, clinical, and disease characteristics of COVID-19-suspected, confirmed, and healthy cases are summarized in Table 1. We observed the significant difference (p < 0.001) in the frequency rate of MS types between groups. The frequency rate of suspected cases had RRMS 57 (87.7%), followed by PPMS 5 (7.7%) and CIS 2(3.1%). In the confirmed group 3 (37.5%) had RRMS and 4 (50%) had PPMS.

Additionally, we found significant differences in frequency rate of MS patients’ comorbid condition such as hypo and hyperthyroidism, cardiovascular, and pulmonary disease among the 3 groups. Moreover, we found that in the confirmed group, MS patients with history of high blood pressure, diabetes, and CVD had the same frequency rate (12.5%).

Another significant difference was detected in frequency rate of MS patients who used corticosteroid and CVD drugs in the 3 groups (p = 0.005 and p = 0.04) respectively. 25% of patients in the confirmed group use corticosteroid drug.

62.5% of confirmed cases had disability score 3 or more. These patients need assistance to walk. Moreover, those who were bed or wheelchair increased significantly in the confirmed group (25% and 37.5% respectively).

There were no significant differences between healthy, suspected, and confirmed cases in demographic information, DMT types and, duration of treatment. 62.5% of patients in confirmed group and 70.6% in the suspected group were female. The average age of MS patients in the confirmed group was higher than the mean age of the healthy and suspected groups, but it was not statistically significant (p = 0.2). In addition, our results showed that most of the patients (36.8%) in the suspected group were on IFN β-1a while patients treated with teriflunomide had the lowest chance of being in the COVID-19 suspected group (1.5%). 50% of confirmed cases treated with rituximab.

Additional results showed 50% of confirmed cases and 35.3% of suspected cases had fever (p < 0.001) and most patients in the suspected group reported sore throat (43.8 %) or dry cough (39.7%). History of contact with a case with fever or cough or shortness of breath showed significant difference in the 3 groups (p < 0.001).

## Discussion

In this study, we evaluated COVID-19 frequency and severity among MS patients in the first pandemic wave in Fars province. According to the prior reports, the incidence rate of respiratory infections for MS patients may be higher than the normal population. In addition, the treatment with second lines showed higher rate of infection than those treated with injectable first lines (2, 3, 19). In this regard, it is probable that the prevalence of COVID-19 in MS patients could be more than the normal population. But, the recent studies showed that the frequency rate of COVID-19 infection in MS patients was not different from the incidence of COVID-19 in the general population. In our study, only 5 % of all respondents showed signs of the COVID-19 infection. This rate was similar to the prevalence of COVID-19 in the general population (20). 5 (0.36%) of all MS patients due to the severity of pulmonary symptoms and shortness of breath with positive PCR test or CT required hospital admission. Two of them had critical situation in ICU and eventually died. The other outcome of COVID-19 infection in Italian MS patients was reported by Sormani et al, who was categorized symptomatic MS patients as mild (96%), sever 2%, and critical 6 (3%) of all respondents (one of the critical patients recovered and five died) (12). Fan et al. reported of 1804 MS patients with DMT none had COVID-19 infection(13). An Iranian study group in Tehran reported mild to moderate output of COVID-19 infection in MS patients. Only two cases needed hospitalization without ICU care and they showed the susceptibility may increase in patients who were on B-cell depleting agents (11). Sahraian et al. reported the frequency rate of COVID-19 infection in MS patients is not higher than general population, however, 25% of confirmed cases need to hospitalization which is far more than the general population(21).

Most of MS patients in our study were relatively young females who had low risk job for covid-19 infection (95%). Data collected from many countries suggested that men have a higher risk of severe illness and death (22). As it was reported in the other study, we think the self-isolation and social distancing guidelines stringently are followed by MS patients. These patients had a good knowledge about this viral infection and its prevalence. It is caused experience a lower incidence of COVID-19 infection compared to the general population (23).

We found that 2 (100%) of critical cases or 25% of confirmed cases died. Moreover, Sormani et al reported 5 (83.3%) of critical cases or 8.77% of confirmed cases in ICU died. However the fatality rate of general population with positive test (n=440) in Fars province has been reported 13 cases (2.95%) during one month of first pandemic wave (24). In this survey, the high frequency of mortality rate in confirmed MS patients may be related to some influencing factors like the low number of PCR tests which performed in MS patients in the first pandemic wave and comorbid conditions like diabetes and hypertension. Previous studies suggested that risk of hospitalization and mortality rate increased in MS patients with comorbid conditions (25, 26).

In our study, two patients who died had PPMS and were on rituximab. One of them was bed-restricted and another could walk less than 500 meters. Results showed patients who were on a rituximab, as compared to patients on other DMTs, had a higher frequency for being categorized in the COVID-19 confirmed group 4 (50%). In our study, two patients who needed ICU care and intubation were on rituximab and eventually died. Safavi et al. (2020) reported only two suspected patients required hospitalizations while both of them were on a B cell depleting agent (Rituximab, Ocrelizumab) and eventually both recovered (11). This result is in agreement with Sormani and Quinti’s (2020) findings (12, 27). Increase in the risk of COVID-19 through rituximab therapies was reported also by Sahraian et al. (21). In current study and Sahraian’s report, two MS patients who died were rituximab users also had underlying diseases. However, further studies are needed to understand the role of rituximab in the deaths of these patients. Rituximab and ocrelizumab can increase the risk of infections by elimination a large portion of circulating B-cells and impair the humoral immune response (2). Therefore, MS patients treated with these agents might be more susceptible to COVID-19 infection.

According to previous report, antiviral properties of IFN-β1 may diminish COVID-19 severity (28). It is possible that patients who were treated with IFN-β1 were protected from the infection. Our results showed that, 36.8% of suspected cases and 25% of the confirmed group were on IFN-β1 eventually recovered well from COVID-19 infection.

The MS type was also detected to be associated with the risk of developing COVID-19, and its frequency rate was significantly higher among PPMS patients. Although, these patients may not have normal physical activity and their presence in the community is low, it seems the frequent closed contacts with symptomatic or confirmed patients was not the main cause of its infection. The high rate of comorbid condition, type of DMT and corticosteroid therapy may increase susceptibility to infection.

### Strengths and Limitations

This study was performed in the first pandemic wave with a large sample size in non-hospitalized MS patients which is a better indicator of the society. However, it has some limitations such as (a) provision the medical information of patients by phone is caused the possibility of intentional or inadvertent errors, (b) over or underestimations of suspected cases due to lack of PCR testing for COVID-19 infection in all of suspected patients or who have had symptoms for more than two weeks, and (c) impossibility of estimating the Expanded Disability Status Scale (EDSS) over the phone.

## Conclusion

The present study showed that rate of developing COVID-19 in MS patients are similar to the general population and the frequency of PPMS type, rituximab, and corticosteroid therapy were higher in the confirmed group.

## Data Availability

All data related to this survey are available

## Acknowledgment

The authors would like to thanks all the staff of the Clinical Neurology Research Center for their cooperation in implementing this project.

